# Investigating spatial variability in COVID-19 pandemic severity across 19 geographic areas, Spain, 2020

**DOI:** 10.1101/2020.04.14.20065524

**Authors:** Sushma Dahal, Kenji Mizumoto, Richard Rothenberg, Gerardo Chowell

## Abstract

**Introduction:** Spain has been disproportionately affected by the COVID-19 pandemic, ranking fifth in the world in terms of both total cases and total deaths due to COVID-19 as of May 20, 2020. Here we derived estimates of pandemic severity and assessed its relationship with socio-demographic and healthcare factors.

**Methods:** We retrieved the daily cumulative numbers of laboratory-confirmed COVID-19 cases and deaths in Spain from February 20, 2020 to May 20, 2020. We used statistical methods to estimate the time-delay adjusted case fatality risk (aCFR) for 17 autonomous communities and 2 autonomous cities of Spain. We then assessed how transmission and sociodemographic variables were associated with the aCFR across areas using multivariate regression analysis.

**Results:** We estimated the highest aCFR for Madrid (25.9%) and the average aCFR in Spain (18.2%). Our multivariate regression analysis revealed three statistically significant predictor variables: population size, population density, and the unemployment rate.

**Conclusions:** The estimated aCFR for 10 autonomous communities/cities in Spain are significantly higher than those previously estimated for other geographic regions including China and Korea. Our results suggest that public health interventions focused on densely populated areas and low socioeconomic groups can ameliorate the mortality burden of the COVID-19 pandemic in Spain.

## Introduction

Since the emergence of COVID-19 in Wuhan, China in December 2019, the novel coronavirus (SARS-CoV-2) continues to spread throughout the world, straining and overloading healthcare systems and causing substantial morbidity and mortality burden during a short time period. As of May 20 2020, 4,789,205 confirmed cases including 318,789 deaths attributed to COVID-19 have been reported from 215 countries/territories/areas [1]. Thus far, the US has reported the highest number of cases (30.8%) and deaths (28.0%) globally. Other countries that follow the US in terms of COVID-19 death toll include the United Kingdom (11.1%), Italy (10.1%), France (8.8%) and Spain (8.7%) [1].

The case fatality risk is a useful metric to assess pandemic severity, which is typically estimated as the proportion of deaths among the total number of cases attributed to the disease [2, 3]. However, during the course of outbreak of an infectious disease such as COVID-19, real-time estimates of CFR need to be derived carefully since it is prone to ascertainment bias and right censoring [4, 5]. In particular, the disease spectrum for COVID-19 ranges from asymptomatic and mild infections to severe cases that require hospitalization and specialized supportive care. This may lead to overestimation of the CFR among ascertained cases. On the other hand, there is a delay from illness onset to death for severe cases [6], which could lead to an underestimation of the CFR [5, 7]. Therefore, statistical methods that help mitigate inherent biases in estimates of the CFR should be employed to accurately plan for medical resources such as ICU units and ventilators, which are essential resources to save the lives of critically ill patients [8–10].

Several studies have reported CFR estimates for COVID-19 [11–13]. Overall, these estimates have varied substantially across geographic regions even within the same country. For example, a recent study estimated the time-delay-adjusted CFR at 12.2% for the ground zero of the COVID-19 pandemic: the city of Wuhan [6], whereas for the most affected region in Italy (Lombardia), the delay-adjusted CFR reached 24.7% [14]. The drivers behind the geographical variations in the severity of the COVID-19 pandemic are yet to be investigated but could provide critical information to mitigate the morbidity and mortality impact of this and future pandemics [15].

In this study we aim to estimate the severity of COVID-19 pandemic across 19 geographic areas in Spain and aim to explain how these estimates varied geographically as a function of underlying factors. For the real-time estimation of severity, we adjust for right censoring using established methods [16, 17] and report the estimates of the time-delay adjusted CFR of COVID-19 for 17 autonomous communities (Comunidades Autonomas, CCAA), and 2 autonomous cities of Spain as well as for the entire Spain. We then assessed the association between different transmission and socio-demographic factors and the estimated CFRs across areas using multivariate regression analyses.

## Methods

### Study setting

Spain is situated on the Iberian Peninsula and is divided into 17 autonomous communities (CCAA) and 2 African autonomous cities [18]. Ceuta and Melilla are the 2 African autonomous cities whereas the 17 CCAA include: Andalucía, Aragon, Asturias, Balears, Canarias, Cantabria, Castilla-La Mancha, Castilla y Leon, Catalunya, C. Valenciana, Extremadura, Galicia, Madrid, Murcia, Navarra, Pais Vasco, La Rioja [18].

### Initial cases of COVID-19 in Spain

The first case of COVID-19 in Spain was confirmed on 31st January, 2020 in La Gomera, Canary Islands in a person who was in contact with an infected person while in Germany [19]. By February 27, there were total 12 cases which increased to 45 cases in March 1 and then continued to rise rapidly throughout the country [20]. The incidence trajectory continued to rise for about 4 weeks before a gradual decline. The daily reported number of new cases has not exceeded 1000 for 10 consecutive days since May 11, 2020 [21]

### Data Sources

The Ministry of Health of Spain releases daily reports on COVID-19 cases and deaths [21]. From these reports we retrieved the daily cumulative numbers of reported laboratory-confirmed COVID-19 cases and deaths from February 20, 2020 to May 20, 2020. We then stratified the data into 20 groups that included 17 CCAA, 2 African autonomous cities and for the entire Spain.

For each CCAA we obtained data on demographic, socio-demographic, and healthcare factors from the Statistics National Institute (Instituto Nacional de Estadística) [22] and the annual report of the national health system [23]. The demographic factors we included were: total population size, population density, proportion of population size by region, proportion of the population older than 60 years, proportion of the population older than 70 years, and life expectancy at birth. We included four socio-demographic factors: poverty risk rate, proportion of population of low social class, unemployment rate, and infant mortality rate. To assess the effect of healthcare factors, we obtained the data on percentage of total consolidated expenditure on hospital and specialized services, and public health expenditure per GDP by CCAA from the annual report issued by the national health system, 2018 [23]. Finally, we also included three factors related to transmission dynamics: The number of days from the date the first case was reported to the day the cumulative number cases surpassed one hundred cases, the cumulative morbidity (total number of reported cases) and the cumulative morbidity rate calculated by the cumulative number cases divided by the local population size. Table S1 summarizes the range and median of above variables for 17 CCAA regions in Spain. Additionally, we obtained the shapefiles of the autonomous communities of Spain from the national geographic information system of Spain [24].

### Time-delay adjusted CFR estimation

The crude CFR is defined as the number of cumulative deaths divided by the number of cumulative cases at a specific point in time. For the estimation of CFR in real time during the course of outbreak, we employed the delay from hospitalization to death, h_*s*_, which is assumed to be given by h_*s*_ = H(s) – H(s-1) for s>0 where H(s) is a cumulative density function of the delay from hospitalization to death and follows a gamma distribution with mean 10.1 days and SD 5.4 days, obtained from the previously published paper [6]. Let π_*a,ti*_ be the time-delay adjusted case fatality risk on reported day t_*i*_ in area a, the likelihood function of the estimate π_*a,ti*_ is

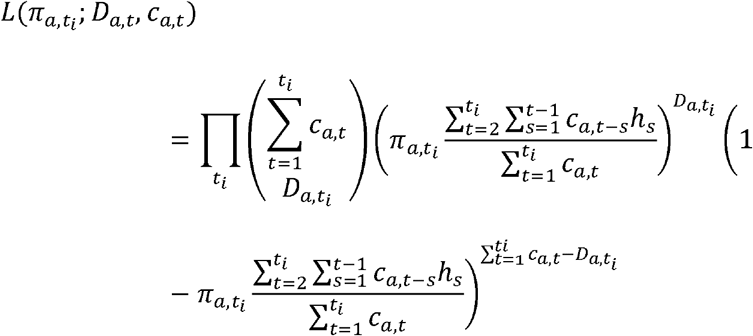

where c_*a,t*_ represents the number of new cases with reported day t in area a, and D_*a,ti*_ is the cumulative number of deaths until reported day t_*i*_ in area *a* [16, 17]. Second parenthesis corresponds to the estimated crude case fatality risk at day t_*i*_ in area a. Among the cumulative cases with reported day *t* in area a, D_*a,ti*_ have died and the remainder have survived the infection. The contribution of those who have died with biased death risk is shown in the middle parenthetical term and the contribution of survivors is presented in the right parenthetical term. We assume that D_*a,ti*_ is the result of the binomial sampling process with probability π_*a,ti*_.

The priors specified in our Bayesian model were non-informative priors. We estimated model parameters using a Monte Carlo Markov Chain (MCMC) method in a Bayesian framework. Posterior distributions of the model parameters were estimated by sampling from the three Markov chains. For each chain, we drew 100,000 samples from the posterior distribution after a burn-in of 20,000 iterations. Convergence of MCMC chains were evaluated using the potential scale reduction statistic [25, 26]. Estimates and 95% credibility intervals for these estimates are based on the posterior probability distribution of each parameter and based on the samples drawn from the posterior distributions.

### Multivariate regression analysis

After quantifying the extent of spatial auto-correlation of CCAA-level time-delay-adjusted CFR median estimates, with the use of Moran’s I statistic with a Delauney triangulation neighbours spatial mixing matrix [27, 28], we explored the association between time-delay-adjusted CFR and predictor variables to identify simplified models with significant factors linked to the variation in CFR estimates across geographic areas in Spain. Multicollinearity tests were examined using variance inflation factors (VIF). Predictors exceeding VIF value of 4.0 were excluded one by one from the multivariate analyses. By fitting all combinations of models given a set of predictors, we derived best minimal models and indicated in Table 2. By taking 1000 draws from the posterior estimates of time-delay-adjusted CFR and running the linear regression 1000 times, the combination of variables with the largest count was selected as the final model. With this model, pooled estimates accounting for both within-model uncertainty and between models uncertainty were derived.

**Table 1.** Summary results of time-delay adjusted case fatality risk of COVID-19 in the 19 areas in Spain, 2020 (As of May 20, 2020)

| Area | Median estimate of time-delayed adjusted case fatality risk as of May 20, 2020 | Range of median estimates of time-delayed adjusted case fatality risk during the study period by geographic region | Crude CFR<sup>‡</sup> |
| --- | --- | --- | --- |
| Spain (National) | 18.2% (95%CrI<sup>§</sup>: 18.0-18.4%) | 12.4-26.8% | 12.0% (95%CI<sup>¶</sup>:11.9-12.1%)<br>27940/233037 |
| Andalucia(AN) | 13.0% (95%CrI: 12.4-13.7%) | 9.8-50.8% | 11.0% (95%CI:10.4-11.5%)<br>1375/12547 |
| Aragon(AR) | 21.3% (95%CrI: 19.9-22.5%) | 15.9-87.5% | 15.2% (95%CrI: 14.2-16.1%)<br>848/5588 |
| Asturias(AS) | 20.3% (95%CrI: 18.3-22.4%) | 3.6-51.7% | 12.9% (95%CrI: 11.6-14.3%)<br>307/2374 |
| Balears(IB) | 15.1% (95%CrI: 13.2-17.1%) | 9.4-51.8% | 10.9% (95%CrI: 9.6-12.4%)<br>221/2024 |
| Canarias(CN) | 8.4% (95%CrI: 7.2-9.7%) | 6.4-52.5% | 6.7% (95%CrI: 5.7-7.8%)<br>155/2307 |
| Cantabria(CB) | 9.5% (95%CrI: 8.3-10.8%) | 7.4-51.8% | 9.2% (95%CrI: 8.0-10.4%)<br>209/2279 |
| Castilla-La Mancha(CM) | 18.7% (95%CrI: 18.0-19.3%) | 15.4-51.3% | 17.4% (95%CrI: 16.8-18.0%)<br>2919/16789 |
| Castilla y Leon(CL) | 11.9% (95%CrI: 11.4-12.4%) | 9.7-51.2% | 10.5% (95%CrI: 10.1-11.0%)<br>1960/18627 |
| Catalunya(CT) | 14.8% (95%CrI: 14.5-15.2%) | 11.6-40.5% | 10.8% (95%CrI: 10.5-11.0%)<br>6021/55888 |
| Ceuta(CE) | 4.6% (95%CrI:1.7-10.0%) | 4.4-51.8% | 3.4% (95%CrI: 0.9-8.4%)<br>4/119 |
| C.Valenciana(VC) | 19.0% (95%CrI: 18.0-19.9%) | 8.0-32.3% | 12.6% (95%CrI: 12.0-13.2%)<br>1383/10987 |
| Extremadura(EX) | 21.7% (95%CrI: 20.0-23.4%) | 15.8-49.5% | 16.6% (95%CrI: 15.3-18.0%)<br>505/3042 |
| Galicia(GA) | 6.9% (95%CrI: 6.4-7.5%) | 5.1-51.4% | 6.7% (95%CrI: 6.2-7.2%)<br>608/9077 |
| Madrid(MD) | 25.9% (95%CrI: 25.3-26.3%) | 13.9-30.5% | 13.3% (95%CrI:13.1-13.6%)<br>8931/67049 |
| Melilla(ME) | 2.9% (95%CrI: 0.7-7.4%) | 2.4-50.8% | 1.7% (95%CrI: 0.2-5.8%)<br>2/121 |
| Murcia(MC) | 9.9% (95%CrI: 8.3-11.5%) | 3.4-53.7% | 9.5% (95%CrI: 8.1-11.0%)<br>149/1570 |
| Navarra(NC) | 11.3% (95%CrI: 10.3-12.3%) | 6.4-51.8% | 9.7% (95%CrI: 8.9-10.6%)<br>506/5195 |
| Pais Vasco(PV) | 17.4% (95%CrI: 16.6-18.2%) | 10.9-71.8% | 11.0% (95%CrI: 10.5-11.6%)<br>1483/13421 |
| La Rioja(RI) | 11.8% (95%CrI:10.6-13.1%) | 3.9-49.7% | 8.8% (95%CrI: 7.9-9.7%)<br>354/4033 |

<sup>§</sup>CrI: 95% credibility intervals (CrI), <sup>‡</sup> Estimates calculated from exact binomial test, <sup>¶</sup>95%CI: 95% confidence interval

**Table 2.**
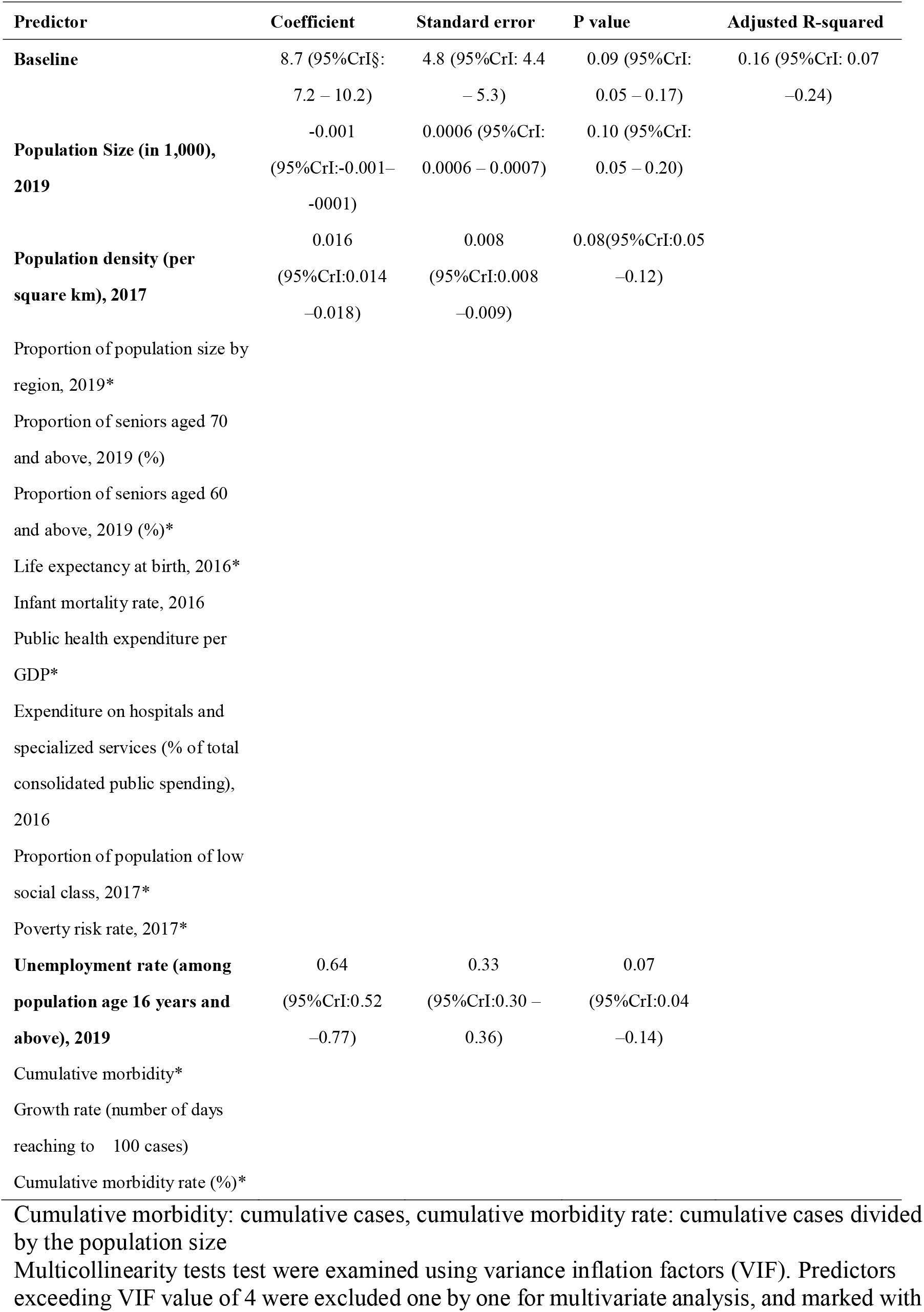
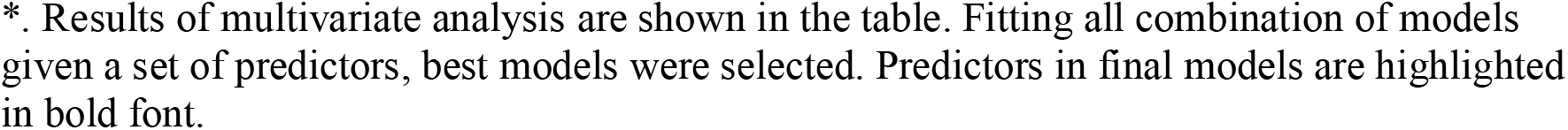
Final multivariate regression model of time-delay-adjusted CFR as a function of socio-demographic variables across autonomous communities of Spain.

| Predictor | Coefficient | Standard error | P value | Adjusted R-squared |
| --- | --- | --- | --- | --- |
| **Baseline** | 8.7 (95%CrI§:<br>7.2 – 10.2) | 4.8 (95%CrI: 4.4<br>– 5.3) | 0.09 (95%CrI:<br>0.05 – 0.17) | 0.16 (95%CrI: 0.07<br>–0.24) |
| **Population Size (in 1,000),**<br>**2019** | -0.001<br>(95%CrI:-0.001–<br>-0001) | 0.0006 (95%CrI:<br>0.0006 – 0.0007) | 0.10 (95%CrI:<br>0.05 – 0.20) |  |
| **Population density (per**<br>**square km), 2017** | 0.016<br>(95%CrI:0.014<br>–0.018) | 0.008<br>(95%CrI:0.008<br>–0.009) | 0.08(95%CrI:0.05<br>–0.12) |  |
| Proportion of population size by<br>region, 2019* |  |  |  |  |
| Proportion of seniors aged 70<br>and above, 2019 (%) |  |  |  |  |
| Proportion of seniors aged 60<br>and above, 2019 (%)* |  |  |  |  |
| Life expectancy at birth, 2016* |  |  |  |  |
| Infant mortality rate, 2016 |  |  |  |  |
| Public health expenditure per<br>GDP* |  |  |  |  |
| Expenditure on hospitals and<br>specialized services (% of total<br>consolidated public spending),<br>2016 |  |  |  |  |
| Proportion of population of low<br>social class, 2017* |  |  |  |  |
| Poverty risk rate, 2017* |  |  |  |  |
| **Unemployment rate (among**<br>**population age 16 years and**<br>**above), 2019** | 0.64<br>(95%CrI:0.52<br>–0.77) | 0.33<br>(95%CrI:0.30 –<br>0.36) | 0.07<br>(95%CrI:0.04<br>–0.14) |  |
| Cumulative morbidity* |  |  |  |  |
| Growth rate (number of days<br>reaching to 100 cases) |  |  |  |  |
| Cumulative morbidity rate (%)* |  |  |  |  |

Cumulative morbidity: cumulative cases, cumulative morbidity rate: cumulative cases divided by the population size
Multicollinearity tests test were examined using variance inflation factors (VIF). Predictors exceeding VIF value of 4 were excluded one by one for multivariate analysis, and marked with
\*. Results of multivariate analysis are shown in the table. Fitting all combination of models given a set of predictors, best models were selected. Predictors in final models are highlighted in bold font.

All statistical analyses were conducted in R version 3.6.1 (R Foundation for Statistical Computing, Vienna, Austria).

## Results

As of May 20, a total of 233,037 cases and 27,940 deaths due to COVID-19 have been reported in Spain. Moreover, the Madrid region has reported the highest number of cases at 67,049 (28.8%) and deaths at 8,931 (32.0%) followed by Catalunya with 55,888 cases (24.0%) and 6,021 deaths (21.5%).

Figure 1 displays the observed and posterior estimates of crude case fatality risk in Spain, 17 CCAAs and 2 autonomous cities (A through T). Day 1 corresponds to March 1st in 2020. Black dots show the crude case fatality risks, and light and dark indicate 95% and 50% credible intervals for posterior estimates, respectively. Our model-based crude CFR fitted the observed data well in all the regions except Aragon where the model did not fit well for first 2 weeks. There was a rapid rise in crude CFR in the Aragon, Asturias, Castilla-La Mancha, C. Valenciana, Extremadura, and in Madrid.

**Fig 1:**
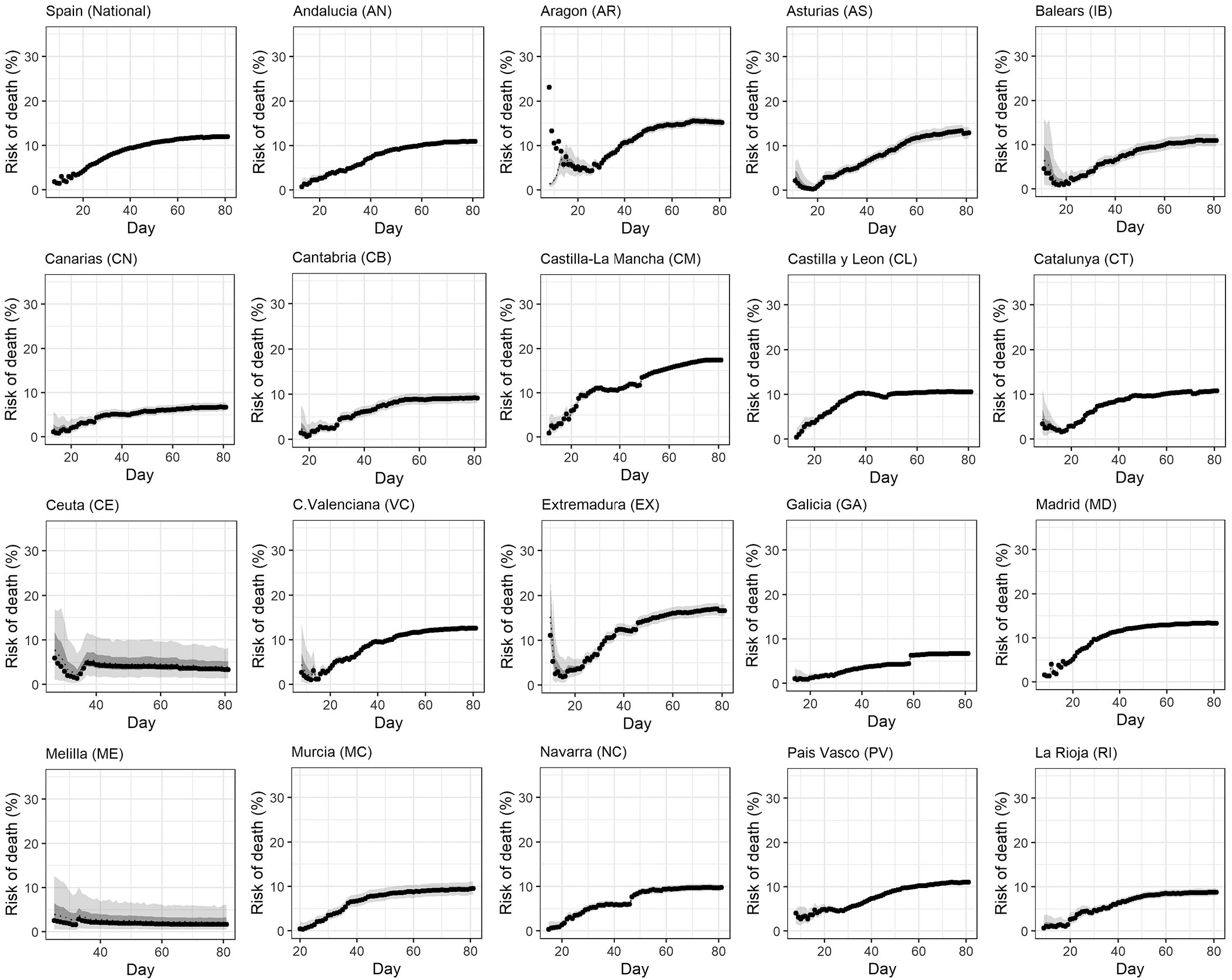
Temporal variation of risk of death caused by COVID-19 by area, Spain, March-May, 2020: crude case fatality risk (cCFR) Observed and posterior estimates from the Bayesian model of crude case fatality risk in (A) Spain (National), (B) Andalucia(AN),(C) Aragon(AR), (D) Asturias(AS),(E) Balears(IB),(F) Canarias(CN),(G) Cantabria(CB),(H) Castilla-La Mancha(CM),(I) Castilla y Leon(CL), (J) Catalunya(CT),(K) Ceuta(CE),(L) C.Valenciana(VC),(M) Extremadura(EX),(N) Galicia(GA),(O) Madrid(MD),(P) Melilla(ME),(Q) Murcia(MC),(R) Navarra(NC),(S) Pais Vasco(PV), and (T) La Rioja(RI). Day 1 corresponds to March 1^st^ in 2020. Black dots show crude case fatality risk, and light and dark indicate 95% and 50% credible intervals for posterior estimates from the Bayesian model, respectively.

Figure 2 illustrates observed and model based posterior estimates of time-delay-adjusted CFR in the 20 areas. Black dots show crude case fatality risks, and light and dark indicate 95% and 50% credible intervals for posterior estimates, respectively. In most of the regions and at the national level, we saw a greater difference between the time delay adjusted CFR and crude CFR in the first 4 weeks of the epidemic followed by a slowly declining difference in the later stage of the epidemic. The graph of the time-delay adjusted CFR varies considerably for different areas. For instance, as the epidemic progressed, the adjusted CFR increased slightly in CN and NC. In contrast, it showed a rapidly increasing trend for first three weeks and then gradually declined in CL. For AR, it was at a high level for the first two weeks after which it started to decline. For the most affected MD region, the time-delay adjusted CFR fluctuated for first 25 days followed by a decline, a leveling off, and then moved upwards.

**Fig 2:**
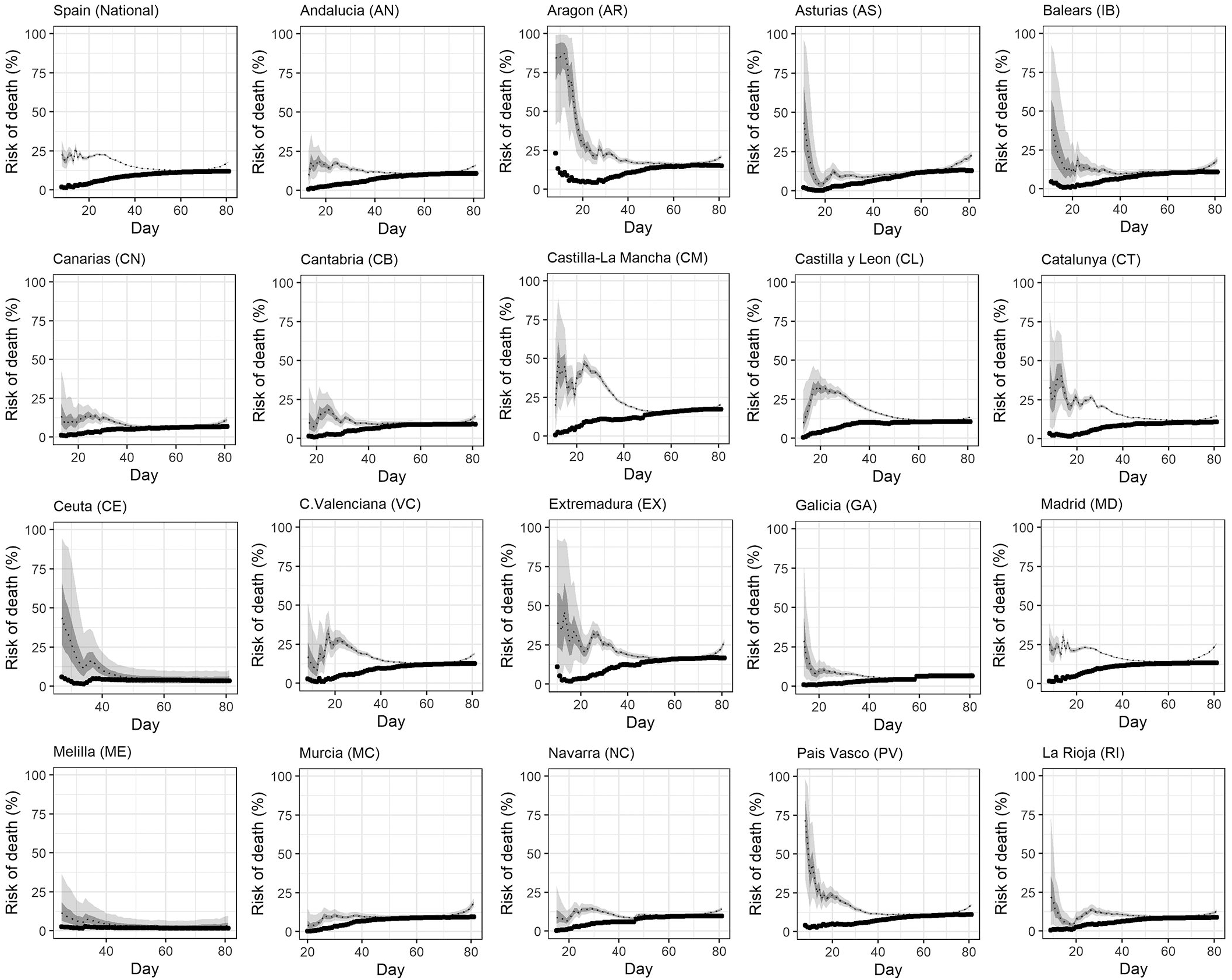
Temporal variation of risk of death caused by COVID-19 by area, Spain, March-May, 2020: time-delay adjusted case fatality risk. Observed and posterior estimates from the Bayesian model of time-delay adjusted case fatality risk in (A) Spain (National), (B) Andalucia(AN),(C) Aragon(AR), (D) Asturias(AS),(E) Balears(IB),(F) Canarias(CN),(G) Cantabria(CB),(H) Castilla-La Mancha(CM),(I) Castilla y Leon(CL), (J) Catalunya(CT),(K) Ceuta(CE),(L) C.Valenciana(VC),(M) Extremadura(EX),(N) Galicia(GA),(O) Madrid(MD),(P) Melilla(ME),(Q) Murcia(MC),(R) Navarra(NC),(S) Pais Vasco(PV), and (T) La Rioja(RI). Day 1 corresponds to March 1^st^ in 2020. Black dots show crude case fatality risk, and light and dark indicate 95% and 50% credible intervals for posterior estimates from the Bayesian model, respectively.

A summary of the time delay adjusted case fatality risk, range of median estimates and crude CFR of COVID-19 across different areas of Spain are presented in Table 1. Our model based posterior estimates of time-delay adjusted CFR are higher than the observed crude CFR. The Madrid autonomous community had the highest time delay adjusted CFR of 25.9% [95% credible interval: 25.3-26.3%] followed by Extremadura (EX) (21.7%) [95%CrI: 20.0-23.4%], Aragon (AR) (21.3%) [95%CrI: 19.9-22.5%], and Asturias(AS) (20.3%) (95%CrI: 18.3-22.4%). The national estimate for Spain was 18.2% (95%CrI: 18.0-18.4%]. Of the 19 autonomous areas of Spain, 13 had the time-delay adjusted CFR greater than 10% (Table 1, figure 3).

**Figure 3.**
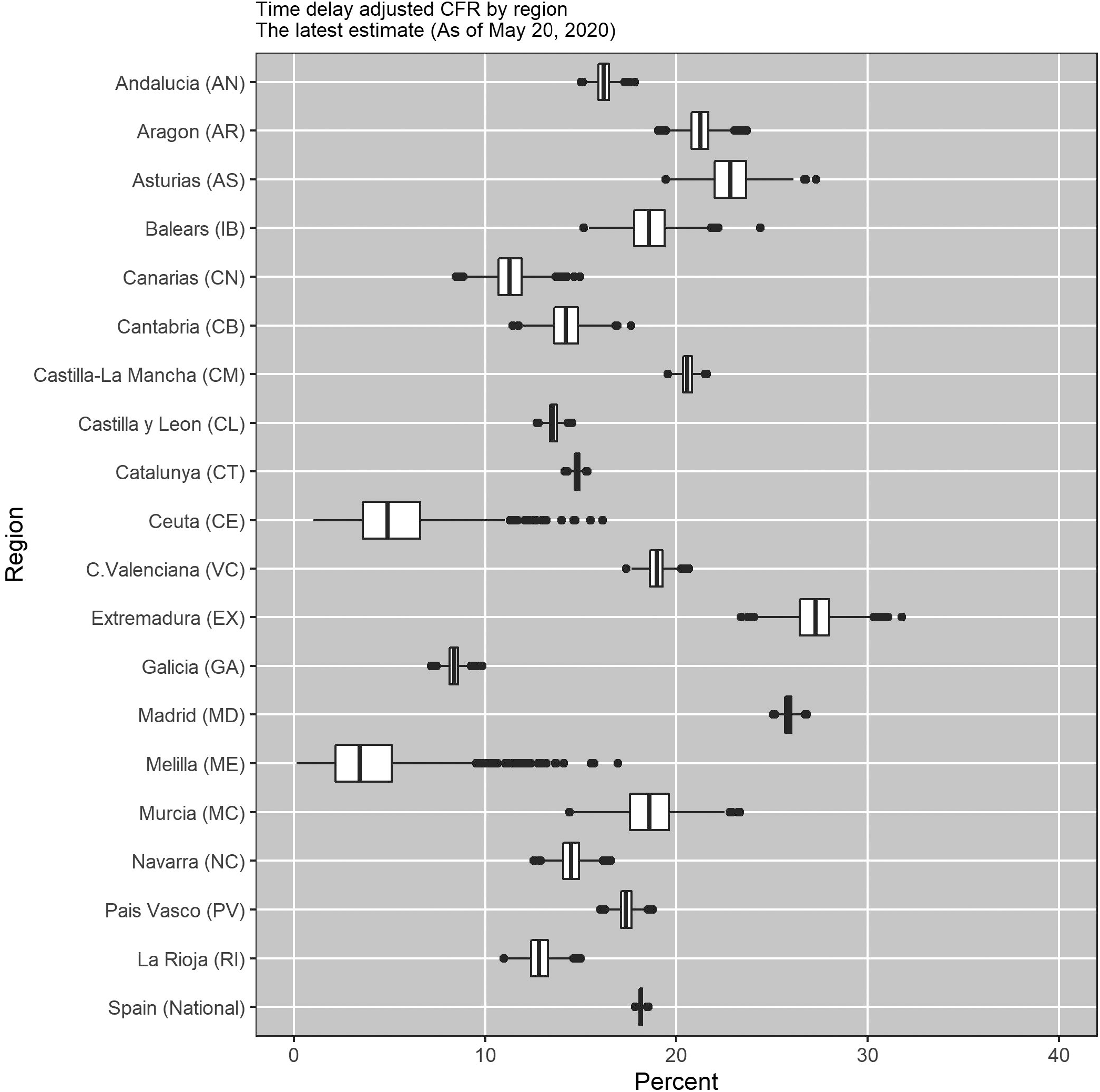
Latest estimates of time-delay adjusted risk of death caused by COVID-19 by area, 2020, Spain (As of May 20, 2020). Distribution of time-delay adjusted case fatality risks derived from the latest estimates (May 20, 2020) are presented. Top to bottom: (A)Spain (National), (B) Andalucia(AN), (C)Aragon(AR), (D) Asturias(AS),(E) Balears(IB), (F) Canarias(CN), (G) Cantabria(CB), (H) Castilla-La Mancha(CM), (I) Castilla y Leon(CL), (J) Catalunya(CT), (K) Ceuta(CE),(L) C.Valenciana(VC),(M) Extremadura(EX),(N) Galicia(GA),(O) Madrid(MD),(P) Melilla(ME),(Q) Murcia(MC),(R) Navarra(NC),(S) Pais Vasco(PV), and (T) La Rioja(RI)

**Figure 4.**
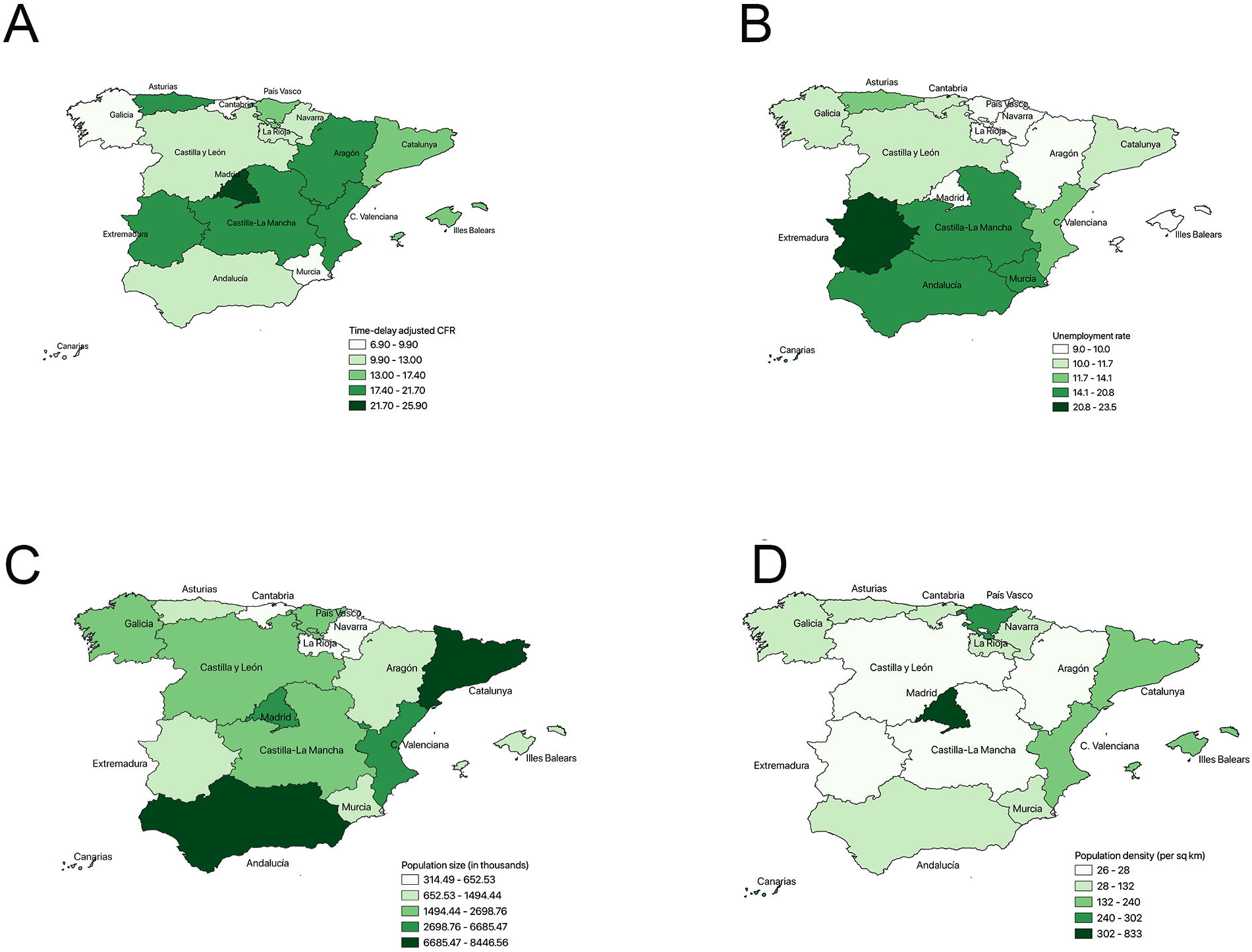
Geographical variability of COVID-19 time-delay adjusted CFR, unemployment rate, population size, and population density across 17 autonomous communities, Spain (As of May 20, 2020). Distribution of time-delay adjusted case fatality risks derived from the latest estimates (May 20, 2020) are presented. Top to b (A) time –delay adjusted case fatality risk as at May 20 2020 (B) Unemployment rate among population aged 16 years and above in 2019 (C) Population size (in thousands) in 2019 (D) Population density (per square km), 2017.

Our analysis of the statistical geospatial heterogeneities in the time-delay adjusted using Moran’s I statistic, which represents the extent of the degree of spatial autocorrelation, was not statistically significant (Moran’s Index = 0.08, P = 0.15). After inspecting VIF, the proportion of population size by region, proportion of seniors aged 60 and above, life expectancy at birth, public health expenditure per GDP, proportion of low social class, poverty risk rate, cumulative morbidity, and the cumulative morbidity rate were excluded from the multivariate analysis.

Autonomous communities with larger population size (p value= 0.10 (95%CrI: 0.05 – 0.20)), higher population density (p value= 0.08 (95%CrI:0.05 – 0.12)) and with a higher unemployment rate (p value=0.07 (95%CrI: 0.04 – 0.14)) experienced higher time-delay adjusted CFRs. These three significant factors explained 16% (95%CrI: 7 – 24)) (adjusted R-squared) of the variation of the pandemic severity across CCAAs (TablelJ2). The correlation matrix of predictors with correlation coefficient (r) is presented in Figure S3. We found that the population size is strongly correlated (r >= 0.7 or r <= −0.7) with the proportion of population size by region (r = 1.00) and the cumulative morbidity while population density is strongly correlated with the proportion of low social class (r = −0.77) and the cumulative morbidity (r = 0.70). Meanwhile the unemployment rate is strongly correlated with the poverty risk rate (r=0.91) and the life expectancy at birth (r=−0.76).

The range of variables included in the linear regression analysis are provided in supplementary Table S1.

## Discussion

In this paper, we have estimated the time delay adjusted case fatality risk of COVID-19 for 19 autonomous areas/cities of Spain. Our estimate of time-delay adjusted CFR in Spain was at 18.2%, but it varied widely across the 19 Spanish areas, with some areas exhibiting higher CFR values such as in Madrid (25.9%), and Extremadura (21.7%) while other areas such as Melilla (2.9%), Ceuta (4.6%), and Galicia (6.9%) experiencing relatively lower CFR values. Of the 19 areas, 13 experienced an adjusted CFR greater than 10%. We also observed a significant positive association of the time-delay adjusted CFR with population size, population density, and unemployment rate across 17 Spanish areas. Our findings suggest the need for additional control efforts as well as generating real time regional severity estimates particularly for areas with high population density and unemployment rate, which have been hit hard by the COVID-19 pandemic.

The adjusted CFR estimates in Spain (18.2%) is higher than the estimates for Wuhan (12.2%)[6], and Korea (1.4%) [29] and Italy (17.4%) [14]. All of these studies employed same methods and hence are comparable. When we compare the estimates for the most affected areas across different countries, the risk in Madrid in Spain (25.9%) is higher than that estimated for Wuhan in China (12.2%) [6], Daegu in Korea (2.4%) [29] and Lombardia in Italy (24.7%) [14]. This difference across countries and regions may be partly explained by differences in population age structure, density, other socio-demographic factors, and the scale of the pandemic in different areas. The median age in Spain (44.9 years) [30] is comparable to that of Italy (45.4 years) but higher than that for China (36.7 years) [31]. Indeed, the elderly population is at risk of severe outcomes from COVID-19 [32–34], which could partly explain the higher severity observed for Italy and Spain. Other factors that could have played a role in these differences may be related to differences in testing strategies. For example, in Korea extensive testing and rigorous contact tracing strategy were implemented [35] while testing prioritized more severe cases in Italy [34] and Spain [36].

Our results from the multivariate analysis found a significant positive association of adjusted CFR with population size and population density, emphasizing the importance of social distancing. We also found a significant positive association between adjusted CFR and unemployment rate across areas in Spain. Unemployment rate is not only an indicator of performance of labor market but also an indicator of socioeconomic status for an area, and this is supported by the strong positive correlation with poverty risk and negative correlation with the life expectancy at birth. Unemployment rate can lead to decreased investment in health, education, nutrition, and good housing condition [37]. In any pandemic situation like COVID-19, the poorer tend to exhibit the highest morbidity and mortality rates. For instance, lower socioeconomic groups were also disproportionately affected by the 1918 influenza pandemic [38, 39]. Data from the US and the UK have shown marked ethnic and racial disparities in COVID-19 death rates [40]. Moreover, preliminary COVID-19 mortality data from the US also indicates a 2-fold age-adjusted death rate among Hispanic/Latino and 1.9-fold among Black/African American compared to Whites [41]. However, we do not know whether the association between unemployment rate and adjusted CFR is inflated by a limited access to COVID-19 testing among low socio-economic groups.

In our study we saw considerable variations in CFR trend across areas. For instance, as the epidemic progressed, the adjusted CFR showed a slightly upward trend in CN and NC, a rapid upward trend followed by the slow decline in CL, and a downward trend in AR. Likewise, for CM the graph showed an upward trend followed by a relative decline and then again an upward trend before declining gradually. The CFR trend for the 19 autonomous areas can be helpful in the planning and implementation of health care services and prevention measures separately for each of them. The downward trend in CFR as seen in some of the areas in our study suggest the improvement in epidemiologic surveillance leading to the increased capture of mild or asymptomatic cases. A higher number of mild and asymptomatic cases also indicate an increase in human-to-human transmission leading to a prolonged epidemic which can be controlled through effective social distancing measures until an effective vaccine or treatment becomes available [6].

The upward trend in CFR indicates that the temporal disease burden exceeded the capacity of healthcare facilities and the surveillance system probably missed many cases during the early phase of the epidemic [6], particularly due to a significant presence of mild and asymptomatic cases. It has been found that about 18% of the COVID-19 infections in Diamond Princess Cruise ship were asymptomatic [42]. The increasing trend in CFR could further be explained the nosocomial transmission affecting the health care workers, inpatients and their visitors [6]. In China, of 44672 confirmed COVID-19 cases, 3.8% was among the health care personnel [43]. Similarly, Wang et al. in their study suspected 41% of the patients to have human-to-human hospital associated transmission of COVID-19 [44].

Our study has some limitations. The preferential ascertainment of severe cases bias in COVID-19 may have spuriously increased our estimate of CFR [5], which is a frequent caveat in this type of studies [45, 46]. Similarly, given the long infection-death time for COVID-19 which ranges between 2 to 8 weeks [30], our estimate may have been affected by delayed reporting bias [5, 7]. Similarly, in our data, the date of report reflects the date of reporting and not the date of onset of illness. Finally, we assumed infant mortality and poverty risk rate as a proxy for areas with low socio-economic groups.

## Conclusion

The risk of death due to COVID-19 in Spain was estimated at 18.2%, but estimates varied substantially across 19 geographic areas. The CFR was as high as 25.9% in Madrid and as low as 2.9% in Melilla. Of the 19 autonomous areas/cities, 13 had a time-delay-adjusted CFR greater than 10% reflecting a disproportionate severity burden of COVID-19 in Spain. Importantly, our estimate of CFR for the most affected Madrid region is higher than previous estimates for the most affected areas within China, Korea, and Italy. Our findings suggest a significant association between unemployment rate, population density, and population size with an increased risk of death due to COVID-19. Further studies with patient level data on mortality, and risk factors could provide a more detailed understanding of the factors shaping the risk of death related to COVID-19.

## Data Availability

The datasets generated during and/or analysed during the current study are available from the corresponding author on reasonable request.

## Note

### Financial support

KM acknowledges support from the Japan Society for the Promotion of Science (JSPS) KAKENHI Grant Number 20H03940, and from the Leading Initiative for Excellent Young Researchers from the Ministry of Education, Culture, Sport, Science & Technology of Japan. GC acknowledges support from NSF grant 1414374 as part of the joint NSF-NIH-USDA Ecology and Evolution of Infectious Diseases program; UK Biotechnology and Biological Sciences Research Council grant BB/M008894/1.

### Additional files

Additional file 1: Table S1. The range of major socio-demographic, and healthcare variables for 17 regions in Spain used in our analyses (linear regression).

Additional file 2: Figure S1. Daily number of reported COVID-9 cases by region in Spain. Day 1 corresponds to March 1st in 2020.

Additional file 3: Figure S2. Daily number of reported COVID-9 deaths by region in Spain. Day 1 corresponds to March 1st in 2020.

Additional file 4: Figure S3. Correlation matrix of predictors.

